# Simplified Within Host and Dose-response models of SARS-CoV-2

**DOI:** 10.1101/2022.09.20.22279832

**Authors:** Jingsi Xu, Jonathan Carruthers, Thomas Finnie, Ian Hall

## Abstract

Understanding the mechanistic dynamics of transmission is key to designing more targeted and effective interventions to limit the spread of infectious diseases. A well-described within-host model allows explicit simulation of how infectiousness changes over time at an individual level. This can then be coupled with dose-response models to investigate the impact of timing on transmission. We collected and compared a range of within-host models used in previous studies and identified a minimally-complex model that provides suitable within-host dynamics while keeping a reduced number of parameters to allow inference and limit unidentifiability issues. Furthermore, non-dimensionalised models were developed to further overcome the uncertainty in estimates of the size of the susceptible cell population, a common problem in many of these approaches. We will discuss these models, and their fit to data from the human challenge study (see Killingley et al. (2022)) for SARS-CoV-2 and the model selection results, which has been performed using ABC-SMC. The parameter posteriors have then used to simulate viral-load based infectiousness profiles via a range of dose-response models, which illustrate the large variability of the periods of infection window observed for COVID-19.

## 1. Introduction

Since December 2019, the SARS-CoV-2 has spread globally, which has infected over half billion of people and caused over six millions of death as well as caused global economic losses (see eg. Chen et al. (2021), Mottaleb et al. (2020), and You et al. (2020)). Mathematical modeling can play a critical role in understanding the viral dynamics in host and estimating the transmission risk, which will provide insights into preventing the spread.

Within host modelling has been widely used to understand the viral dynamics of SARS-CoV-2 infection at an individual level and has many variants under different scenarios and assumptions. The work of Abuin et al. (2020) considers the simplest SIV model and focuses on the equilibrium points with respect to virus extinction and its relations with the initial conditions. Ke et al. (2020) develops a within host model that describes the one-way transport of virus particles from upper respiratory tracts and to lower respiratory tracts. In the mechanistic model of Challenger et al. (2022), the late immune response is triggered by a large magnitude of infected cells and reaches mature via a series of equations, which does not wane once reaches mature and provides accelerated clearance of infected cells. Goncalves et al. (2021) assumes that the productively infected cells will produce infectious virus and non-infectious virus with probability. Gonçalves et al. (2021) further considers an additional model that incorporates the antigen-dependent immune response, however, this more complicated structure reduces the accuracy of parameter inference despite reduced BIC. In the model of Goyal et al. (2021) (also cf. Goyal et al. (2020)), the infected cells are cleared by early immunity response and late T cell response. The late T cell response is driven by the SARS-CoV-2-specific effort cells raising from two stages of precursors cells. In Goyal et al. (2021), the use of a logistic function for emergence of the adaptive response (with the derived parameters) suggests a relatively fast switch. The SIV system fitted in Goyal Goyal et al. (2020) suggests viral load is quickly removed which suggests the viral load can be seen as a scaling of infected cells. Hence, based on these two observations, we simplify the model in Goyal et al. (2021) in this paper and extend it to two scenarios where the decay of the viral load is driven by the depletion of susceptible cells and adaptive response respectively, which reduces uncertainty in parameter inference and gives more insights into considering the viral shedding. Both models can explain the data from the human challenge study Killingley et al. (2022), which can be seen in Section 4.

A series of studies have considered viral load for SARS-CoV-2 as an important determinant for evaluating infectiousness, disease severity, and transmission risk (see eg. Pujadas et al. (2020), Fajnzylber et al. (2020)). The magnitude of the viral load has been considered as an important metric for evaluating the infectivity and transmission rate in communities via dose response models Watanabe et al. (2010), and patients with higher viral load are closely related Marks et al. (2021). Dose-response models enable us to evaluate the probability of transmission through viral load in a quantitative way. Only two papers, Goyal et al. (2021) and Ke et al. (2020), link viral load and transmission of SARS-CoV-2 infection. Goyal et al. (2021) splits the transmission into contagiousness and infectiousness, which is defined by the Hill function with the same given parameters. Ke et al. (2020) assumes that only a proportion of virus could reach the respiratory tract of exposed contacts and so the probability of transmission is defined as the probability that at least one of viruses causes infection. An exponential form dose-response model with a Michealis-Menten term representing the amount of virus shed from upper respiratory tracts is adopted in Ke et al. (2020). Hence, another goal of this paper is to further study the viral load dependent infectiousness. We investigate two mechanisms, the competing risk framework Haas et al. (2014) (for parameter parsimony choosing 2 comparative models: the basic exponential model and the approximate beta-Poisson model), and the logistic growth framework used in viral load models found in the literature review (eg Goyal et al. (2021)).

## 2. Formulation

The model Goyal et al. (2021) is based on the concept of susceptible cells (*S*), infected cells (*I*) and viral load (*V*) that may be shed or detected. The infected cells are cleared by early immunity response and the late T cell response driven by the SARS-CoV-2-specific effector cells, and the effector cells must first go through a series of maturation steps via precursors cells. Based upon this model, we note that the free virus in the SIV system fitted in Goyal is quickly killed and so essentially *V* is a scaling of *I* state. Equally the logistic function on adaptive response with the derived parameters in the published paper suggests a relatively fast switch and so the system can be simplified to

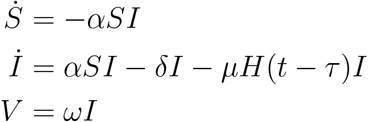

where *τ* is the timescale for adaptive response to become ‘effective’ and *H* is a Heaviside function (so zero when argument is negative and 1 otherwise). In this model, *α* represents the infection rate of susceptible cells *S*. The infected cells are cleared with rate *δ* by early immunity response and by adaptive immunity response with rate *µH*(*t* − *τ*).

Note this is mathematically similar to a SIR compartment epidemic model Keeling & Rohani (2007) (but the R state is not relevant as removal does not affect replication process). *V* is then the free virus that may be emitted (and may be calibrated to the human challenge data Killingley et al. (2022)). If we assume this virus is infectious material then the *V* state can directly be fitted to the PFU dataset. Because PCR data measures DNA we may add a further state to the model

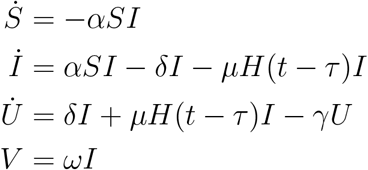

and then we may fit the PCR data to *P* = *θ*(*I* + *U*).

To reduce model complexity we focus on fitting to the PFU data set. Note that this is not readily identifiable as *S* and *I* can be arbitrarily scaled. To avoid this we non-dimensionalise the model such that *S* = *XN, I* = *Y N*, define new parameters *ρ* = *αN/δ, ϕ* = *ωN* for some constant *N* representing the total susceptible cells in the system at time of infection (i.e. *N* = *S*(0) rather than *S*(0) + *I*(0)). Then

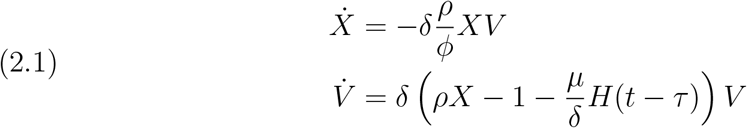

and by definition *X*(0) = 1 and thus we need to identify 6 parameters *ρ, ϕ, δ, µ* and *τ* and the initial condition on *V* (0) = *V*_0_. In model (2.1), *ρ* represents the threshold parameter in the model (so deterministically if *ρ >* 1 the virus grows and if *ρ <* 1 it decays) whilst *ϕ* modifies the removal of susceptible cells (and when *t < τ* it determines the magnitude of the peak viral load for *V*, see supplementary material S.4).

It is important to note that (2.1) despite the the care to non-dimensionalise the system has unidentifiable or rather non-unique parameterisation. Very similar model fits can be derived from a relatively small *ϕ* choice (where we would expect *τ* to be a time greater than the 14 days of observations in the calibration data and so the decay to be due to depletion of susceptible cells and *µ* would have no information either) and a large *ϕ* choice where *τ* would be the time of peak viral load and equivalent to a piece-wise exponential model.

In the scenario of a large susceptible cell population and early immune response boosting then *X* ∼ 1 for the timescales considered (so *ϕ* ≫ *δρ*) then (2.1) becomes

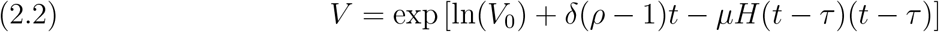

In this situation then the only identifiable parameters are the initial condition (derived from the intercept of a log-linear model), the growth rate (*r*_+_ = *δ*(*ρ* − 1)) and the eventual decay rate after the time of peak viral load *r*_*−*_ = *r*_+_ − *µ*. In order to estimate either *δ* or *ρ* we would require independent estimates of the other parameter.

In the situation that the susceptible cell population size is more limited with later immune response boosting we may consider further analysis given in supplementary material S.4.

### 2.1. Model calibration method

Approximate Bayesian Computation Sequential Monte Carlo (ABC-SMC) algorithm is adopted to conduct parameter inference (cf. Toni et al. (2009), and Minter & Retkute (2019)), in which a multivariate normal distribution with optimal local covariance matrix is used as the perturbation kernel Filippi et al. (2013). For ABC-SMC algorithm, the distance function is defined as:

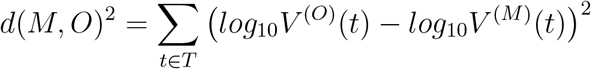

in which *T* represents the set of time points of the data, and *V* ^(*O*)^ and *V* ^(*M*)^ represent the observations and simulated outputs respectively. For each generation of iterations, 250 particles are collected and the number of generations is 10 for improving accuracy.

Three scenarios have been considered during the parameter inference for equation 2.1 and The first scenario assumes the values of *τ* to be the corresponding time of peak viral load for equation 2.2 and we assume that *δ* = 1 in this case. The prior distributions for the rest of the parameters are given by:

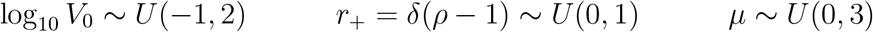

Scenario 2 considers equation 2.1 when *τ* is in between the corresponding time of peak viral load and 14 days, in which *τ* and *µ* are identifiable. Scenario 3 constrains the values of *τ* to be greater than 14 days so that there is no immune boost impact from the parameter *µ*. Apart from that, the prior distributions for the rest parameters are the same:

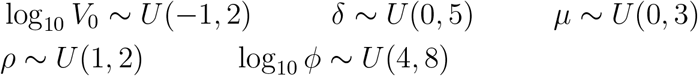

The data from Human Challenge Study has been used to carry out parameter inference Killingley et al. (2022). Posterior distributions and predictions for these three scenarios can be seen in Figure 1 to Figure 6 (and in supplementary material Figures 10 through to 15 respectively).

**Figure 1:**
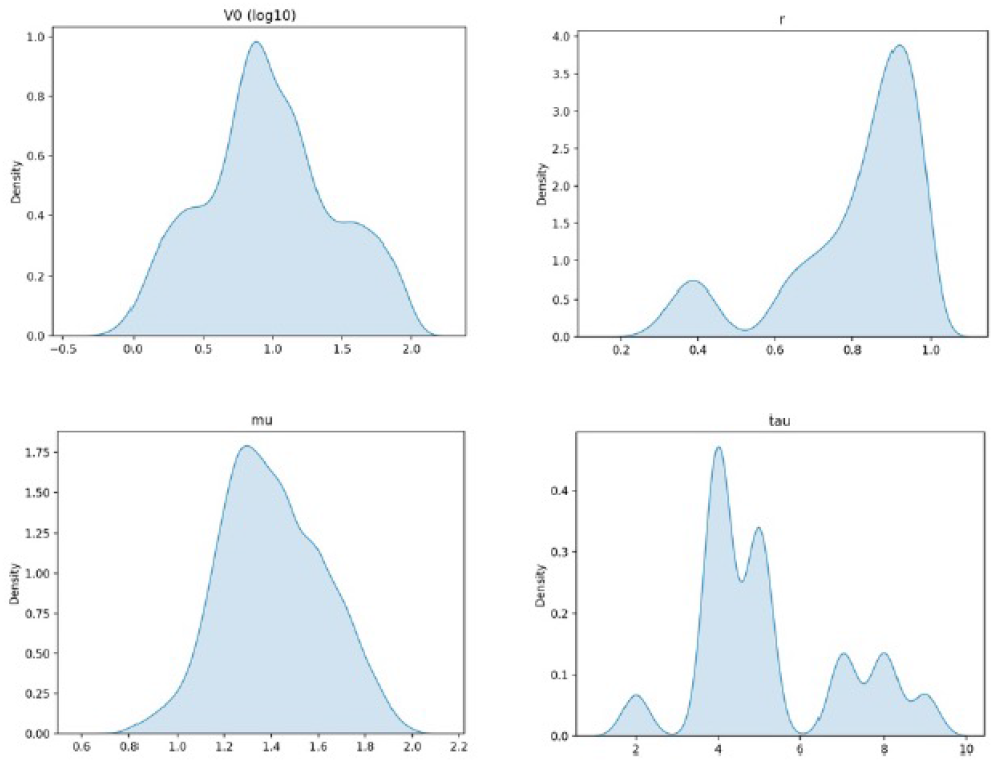
(Scenario 1) Approximate posterior distribution of parameter values from the result of ABC-SMC with model from equation 2.2 using throat data Killingley et al. (2022)

**Figure 2:**
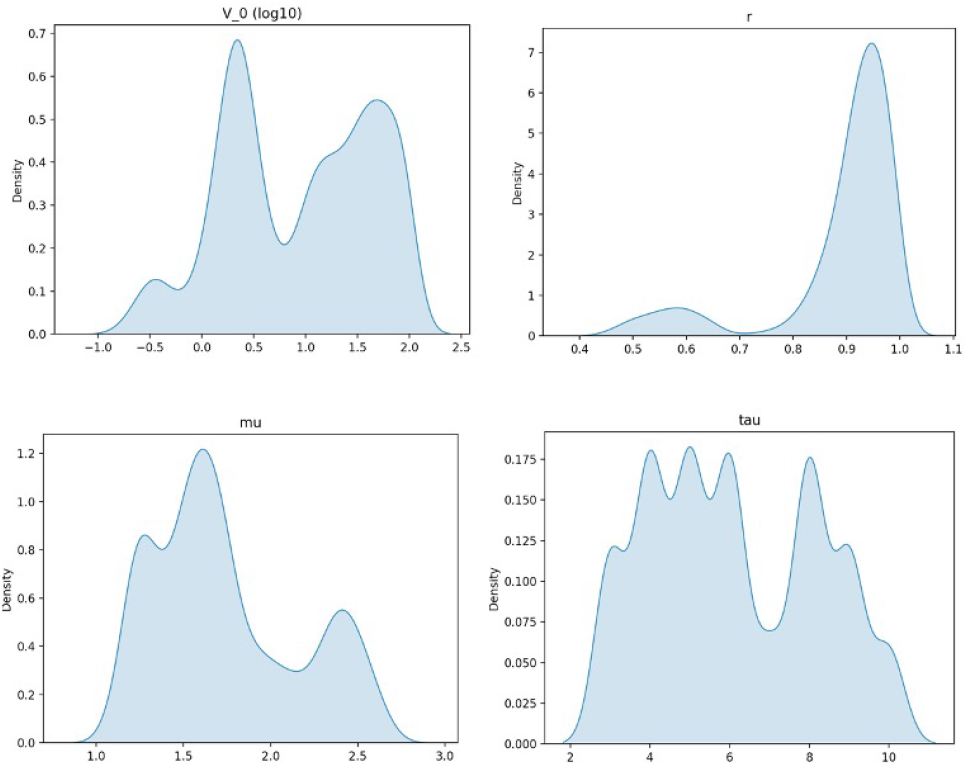
(Scenario 1) Approximate Posterior distribution of parameter values from the result of ABC-SMC with model from equation 2.2 using mid-turbinate data Killingley et al. (2022)

**Figure 3:**
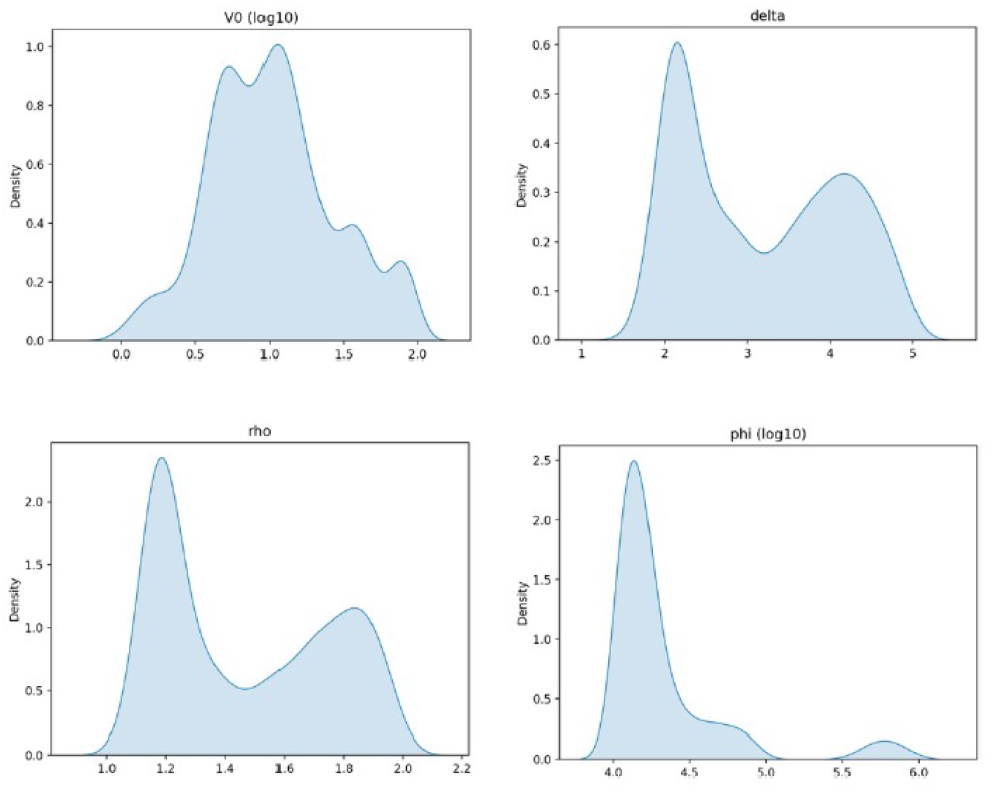
(Scenario 2) Approximate Posterior distribution of parameter values from the result of ABC-SMC with model from equation 2.1 using throat data Killingley et al. (2022)

**Figure 4:**
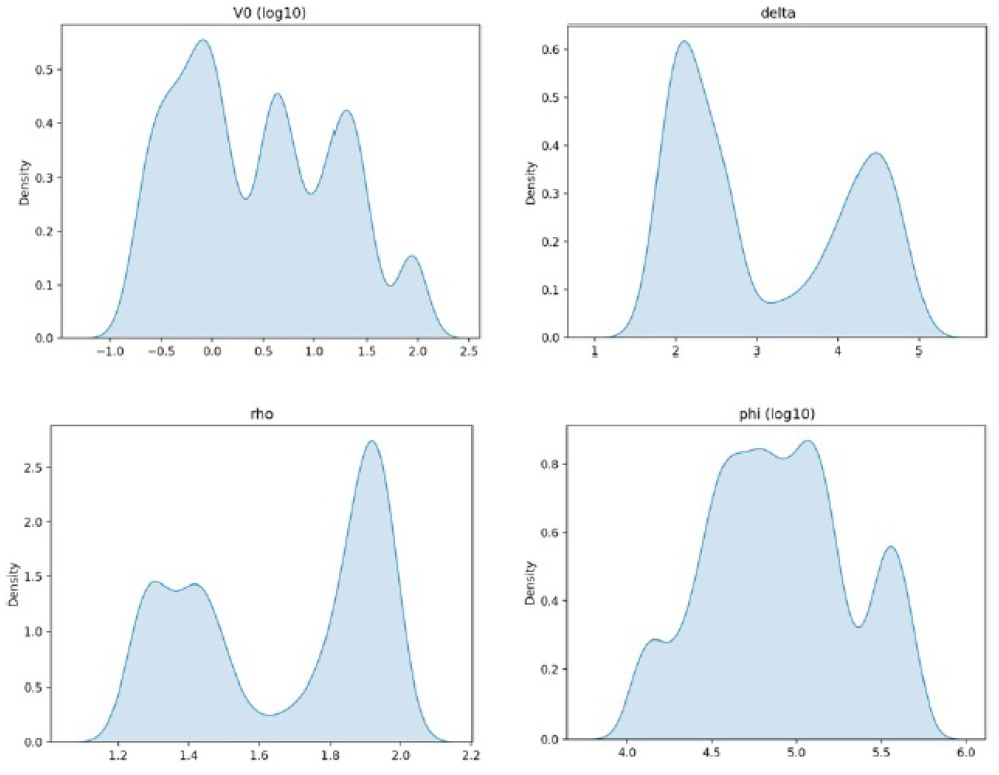
(Scenario 2) Approximate Posterior distribution of parameter values from the result of ABC-SMC with model from equation 2.1 using throat data Killingley et al. (2022)

**Figure 5:**
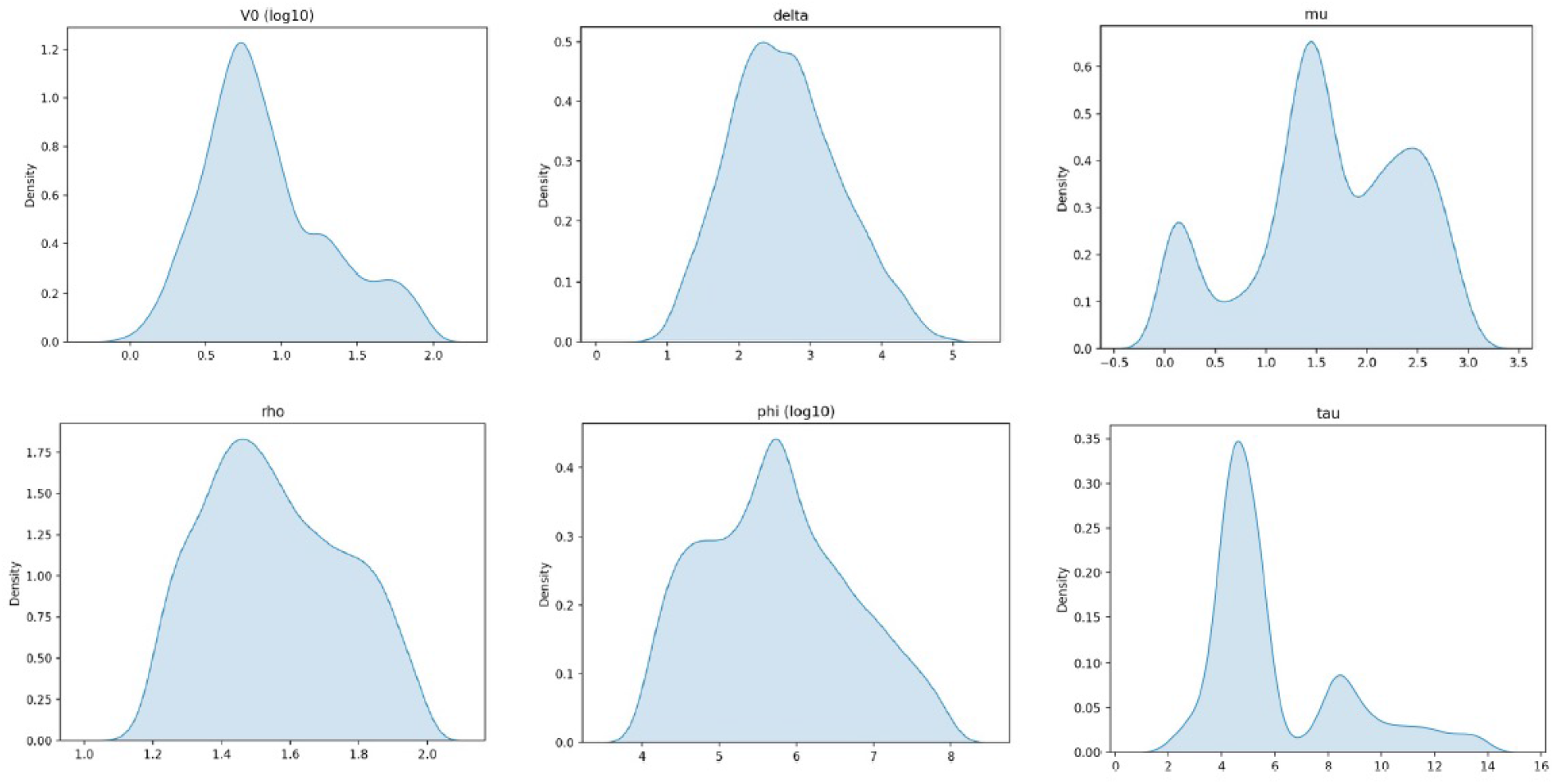
(Scenario 3) Approximate Posterior distribution of parameter values from the result of ABC-SMC with model from equation 2.1 using throat data Killingley et al. (2022)

**Figure 6:**
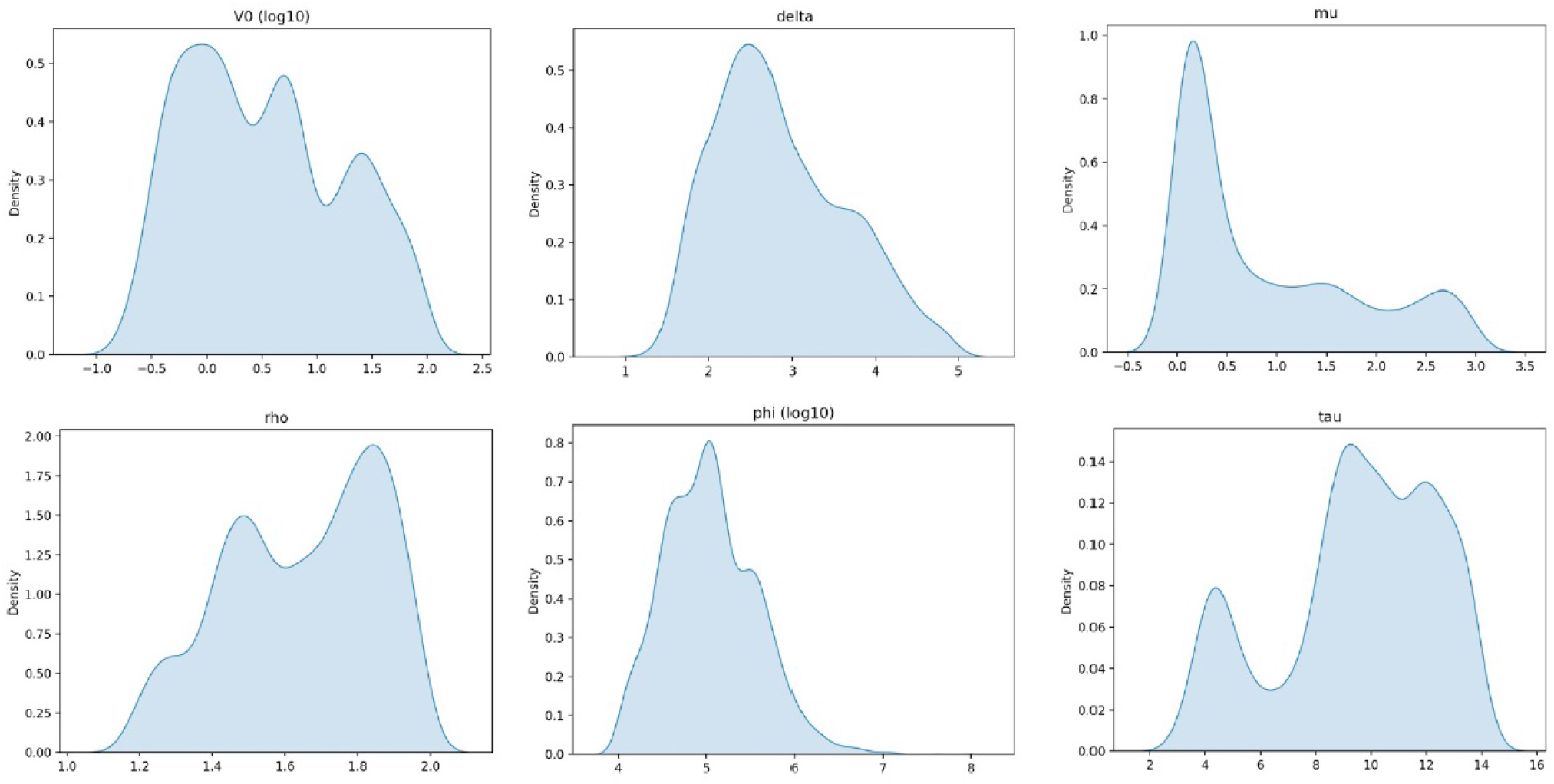
(Scenario 3) Approximate Posterior distribution of parameter values from the result of ABC-SMC with model from equation 2.1 using mid-turbinate data Killingley et al. (2022)

## 3. Dose-response models and probability of infectiousness through different contact routes

Dose response models attempt to describe human challenge data Killingley et al. (2022) and by explicitly stating a mechanism offer an opportunity to extrapolate to other dose regimes. Instead we investigate two mechanisms, the competing risk framework borrowed from bacterial infections Haas et al. (2014) and the logistic growth framework used in viral load models found in literature review Goyal et al. (2021).

Extension to the competing risks model has been investigated in detail elsewhere Pratt et al. (2020), Haas et al. (2014) and here we follow material from Pratt et al. (2020) only for coherence and transparency of report but encourage reader to core material. In a competing risks formulation there are two events that may occur (here establishment of virus or removal). The usual assumption is made that the hazards of establishment and removal are constant (and so the removal and establishment times are Exponentially distributed). In this case the ratio of the mean time to events occurring is given by the parameter, here denoted *γ*.

Deposition to the infection site is a further filter on the exposure from environment. This may be viewed as a Binomial process with probability of deposition *υ* which may vary depending on activity level of the host and particle size inhaled, but will be assumed not to vary between individual virus particles, whilst dose *D* will depend on (atmospheric) dispersion of the agent and so will depend on the individual’s position relative to the source. The probability *Q* of removal of all virus particles without infection occurring is then

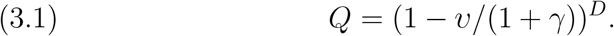

Ordinarily one assumes that the number of virus particles in a unit volume of inhaled air is modelled as a Poisson process Haas et al. (2014), that is we assume that spores are randomly distributed in the atmosphere local to the inhaler and with expected dose proportional to the amount of air inhaled, *D* ∼ Pois(*x*). It is worth reflecting that the infectious dose that would be expected to infect 50% of the population exposed (the median infectious dose, denoted *d*_50_) is a useful metric. In this case we find the dose-response function takes the form of an Exponential function,

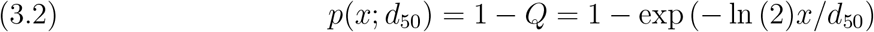

where *d*_50_ = (1 + *γ*)*/*(*υ* ln 2) in notation of derivation. Note that, here and throughout this section, *x* = *βv* and *β* is a scaling factor for viral dose *v* estimated from model (2.1).

We may consider variation at a host level in deposition, response and removal rates which results in alternative dose response curves based on generalisation of Hyper-geometric functions (as described in Pratt et al. (2020) and Abramowitz & Stegun (1972)). In this work we limit ourselves on grounds of parsimony to only consider the approximate Beta Poisson dose response form

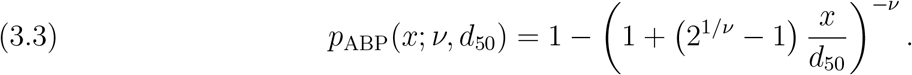

This dose-response relationship is commonly applied in the literature and confusingly also called the Beta-Poisson model Teunis & Havelaar (2000), Huang et al. (2009), Tamrakar et al. (2011), Haas et al. (2014).

An alternative dose response relation was used by Goyal et al Goyal et al. (2021) termed in that work the Hill function but is related to the logistic growth equation commonly used in ecology. Consider a probability of infection *p* that has a maximum attainable probability of 1 and whose rate of change as the dose changes is

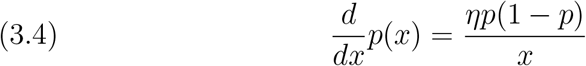

and so

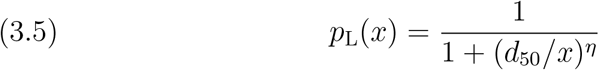

so as *x* → ∞ then *p*_*L*_(*x*) → 1 and when *x* = 0 we have *p*_*L*_(0) = 0.

This model (as with the others presented) will be sigmoid as dose increases but as *η* gets larger the profile is increasingly sharp. A sharp profile can be interpreted as there being effectively a threshold dose above which infection is likely to occur. However a shallow curve may be worse in terms of control as individuals spend longer infectious non-negligible probabilities of infection. This may be worse in terms of infection as spending 4 days with 50% chance of transmission is the same as spending 2 days with close to 100% chance of transmission. We also note that when *η* = *ν* = 1 equations (3.3) and (3.5) are identical.

## 4. Results

### 4.1. Fitting the simplified model to Human Challenge data

The following subsections focus on the resulting posterior distributions arising from the model in various scenarios. The associated visual comparison to the human challenge individual longitudinal data Killingley et al. (2022) itself of each model for each trial participant is given in supplementary material S.

#### 4.1.1. Scenario 1: Fast adaptive immune response, large susceptible pool of cells

In this scenario we fit equation (2.2) to the human challenge data Killingley et al. (2022) for mid-turbinate and throat independently. To simplify the model we define *τ* to be the time of the peak viral load and so uncertainty in other parameters may not be fully realised.

Figures 1 and 2 show the posterior distribution plots for each parameter for throat and midturbinate respectively whilst Figure 10 and Figure 11 show the corresponding model fits in both locations for the 18 volunteers. Note that the posterior distribution plots for each individual volunteer are exhibited in Figure 16 and Figure 17 in supplementary material respectively.

This model has four parameters corresponding to panels in Figures 1 and 2. The top left panel gives the estimated initial number of viral particles *V*_0_, the top right gives the estimated growth rate *r* = *δ*(*ρ* − 1), the bottom left the estimated decay rate *µ* and bottom right is the assigned value of *τ* (given by time of peak reported viral load hence the point estimates rather than distribution).

Note that the nose tends to have faster decay rates than those associated with the throat though some participants have remarkably similar decay rates.

#### 4.1.2. Scenario 2: Slow adaptive immune response, relatively small susceptible pool of cells

In this scenario we fit equation (2.1) to the human challenge data Killingley et al. (2022) for mid-turbinate and throat independently, however, to simplify the model we define *τ* to be greater than the timescale the experiment was conducted over (14 days).

Figures 3 and 4 show the posterior distribution plots for each parameter whilst Figures 13 and 12 show the corresponding model fits in both locations for the 18 volunteers. Note that the posterior distribution plots for each individual volunteer are exhibited in Figures 19 and 18 respectively in supplementary material.

This model also has four parameters corresponding to panels in Figures 3 and 4. The top left panel gives the estimated initial number of viral particles *V*_0_, the top right gives the estimated decay rate *δ*, the bottom left the estimated threshold parameter *ρ* and bottom right is the estimated value of *ϕ* a measure of the number of cells affected (on a log base 10 scale).

#### 4.1.3. Scenario 3: Intermediate adaptive immune response, relatively small susceptible pool of cells

In this scenario we fit equation (2.1) to the human challenge data Killingley et al. (2022) for mid-turbinate and throat independently, however, to ensure a different calibration to the other scenarios we force *τ* to be greater than the peak time and less than or equal to the timescale the experiment was conducted over (14 days).

Figures 5 and 6 show the posterior distribution plots for each parameter for each participant whilst Figures 15 and 14 show the corresponding model fits in both locations for the 18 volunteers. Note that the posterior distribution plots for each individual volunteer are exhibited in 20 and 21 in supplementary material.

This model also has six parameters corresponding to panels in Figures 5 and 6. The top left panel gives the estimated initial number of viral particles *V*_0_ (now on a log base 10 scale), the top central panel gives the estimated decay rate *δ*, the top right panel gives estimates of *µ*, the bottom left the estimated threshold parameter *ρ* and bottom central is the estimated value of *ϕ* a measure of the number of cells affected (also on a log base 10 scale) and finally the bottom right panel the estimate of *τ*.

### 4.2. Results of linking in host model with dose response model

The dose-response models highly rely on the accurate measure of ID50. In the work of Killingley et al. (2022), 53% of patient developed PCR-confirmed infection under the inoculation of 10TCID50 (TCID50 is the median tissue culture infectious dose) of SARS-CoV-2, and 95% confidence interval is (35,70). Hence, in this paper, we use 10TCID50 as an approximate for ID50, which is 50PFU. Here we show results for a range of assumed *β* (this being a scaling factor of dose from infector to infectee, a proxy for distance of time spent together) values for sample from in host parameter estimates in exponential model (3.2), approximate beta-Poisson (3.3), and Hill function (3.5). To illustrate the result, we sample a set of data from the posterior predictions of mid-turbinate data of the case A as an example (see Figure 14), in which the viral load approaches the peak at day 8.

Figure 7 shows the probability of infection for viral scaling factors of 10% (green), 1% (orange), and 0.1% (blue) using equation (3.2). This illustrates a fairly broad infection window of about 8 days for the stronger transfer factor.

**Figure 7:**
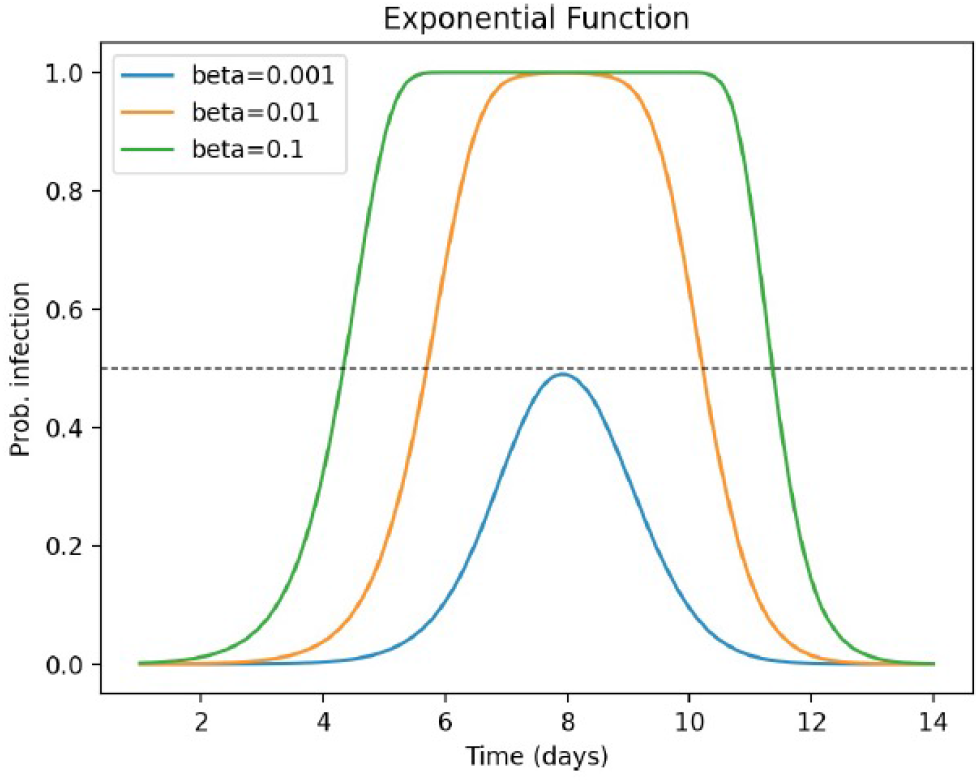
Viral load dependent probability of infection from exponential dose-response function (3.2)

Figure 8 shows the probability of transmission using equation (3.3). Here we have an additional parameter, *ν*, to compare (top left, *ν* = 0.01 ; top right *ν* = 0.1 ; bottom left, *ν* = 1 and bottom right *ν* = 10). For large *ν* the results should tend to those in Figure 7. As *ν* become smaller the infectious period tends to increase, whilst for lower values of *β* changes in *ν* does not tend to change maximal value of the probability of transmission at contact.

**Figure 8:**
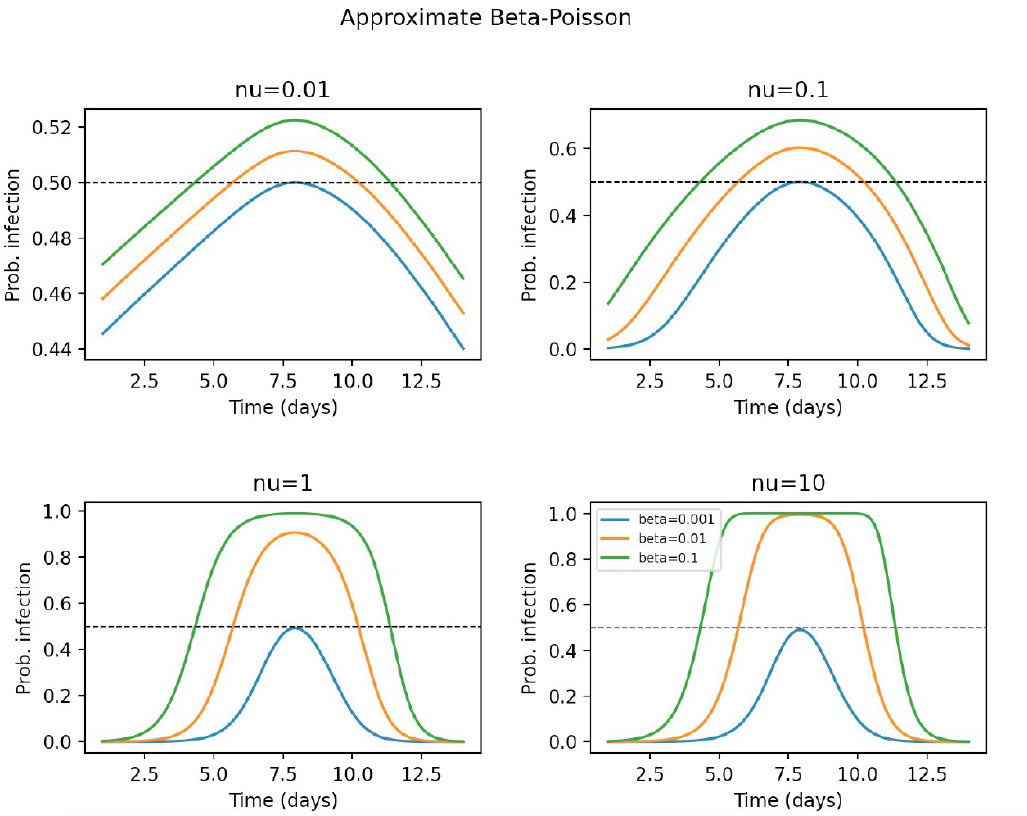
Viral load dependent probability of infection from approximate Beta-Poisson dose-response function (3.3) under different values of *ν*

Figure 9 shows the probability of transmission using equation (3.5). Here we have an additional parameter (*η*) to compare (top left, *η* = 0.01 ; top right *η* = 0.1 ; bottom left, *η* = 1 and bottom right *η* = 10). For *η* = 1 the results are the same as when *ν* = 1 in Figure 8 and for smaller *η* the pattern is similar for smaller *ν* above. For large *η* we see a much more pronounced and shorter infectious period, similar to that reported in Goyal et al. (2021).

**Figure 9:**
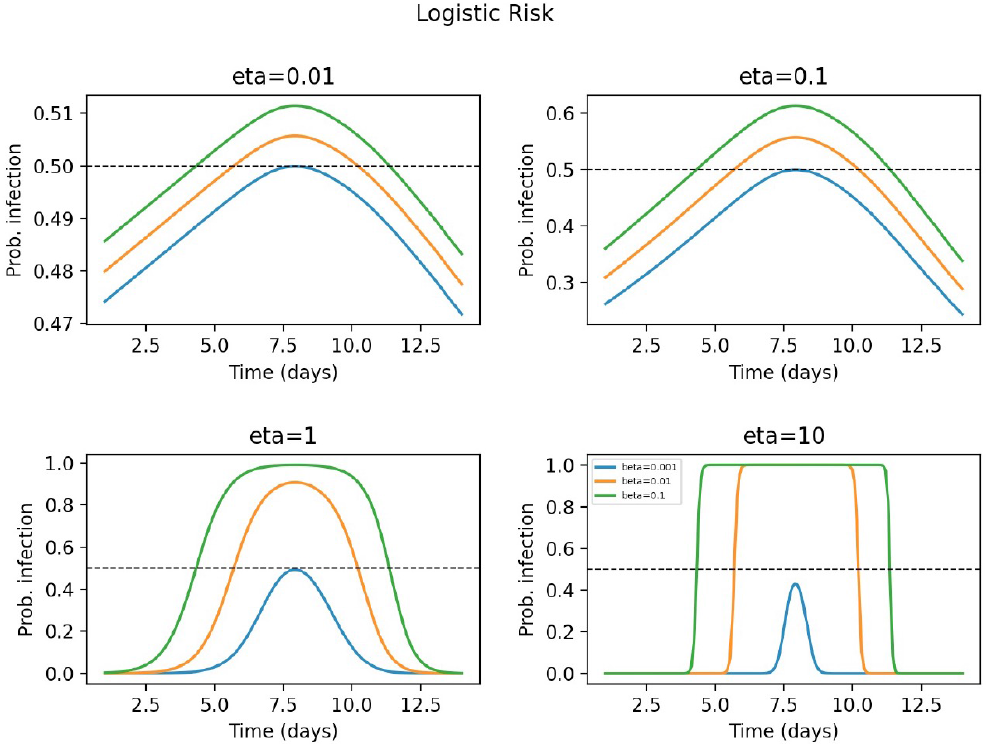
Viral load dependent probability of infection from logistic dose-response function (3.5) under different values of *η*

**Figure 10:**
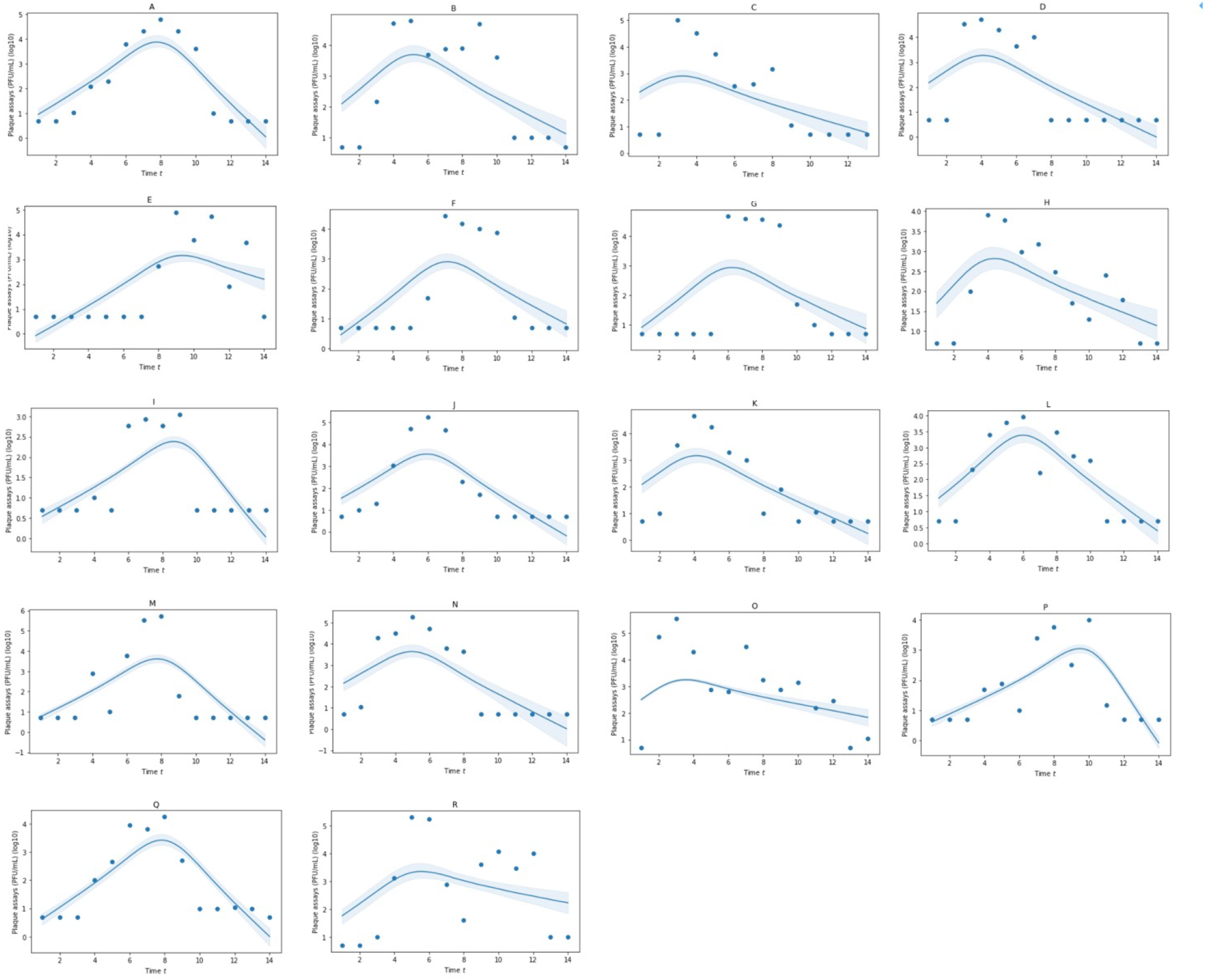
(Scenario 1) Posterior predictions of infectious virus (PFU/mL) from mid-turbinate for 18 participants from the Human Challenge Study Killingley et al. (2022). Model is that derived in equation (2.2). Shaded regions represent the 95% credible interval. Values of *τ* are the corresponding time of peak viral load

**Figure 11:**
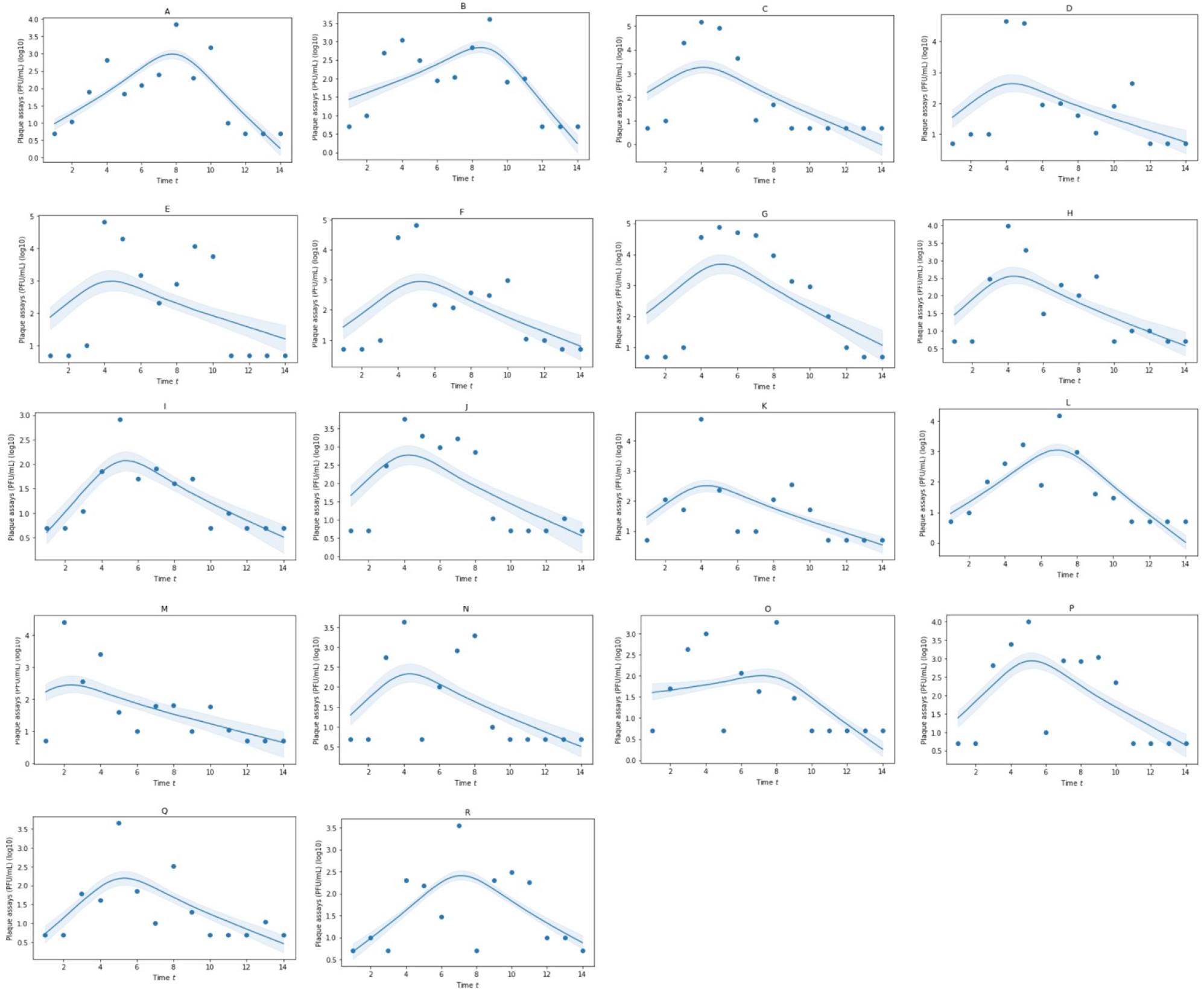
(Scenario 1) Posterior predictions of infectious virus (PFU/mL) from throat for 18 participants from the Human Challenge Study Killingley et al. (2022). Model is that derived in equation (2.2). Shaded regions represent the 95% credible interval. Values of *τ* are the corresponding time of peak viral load

**Figure 12:**
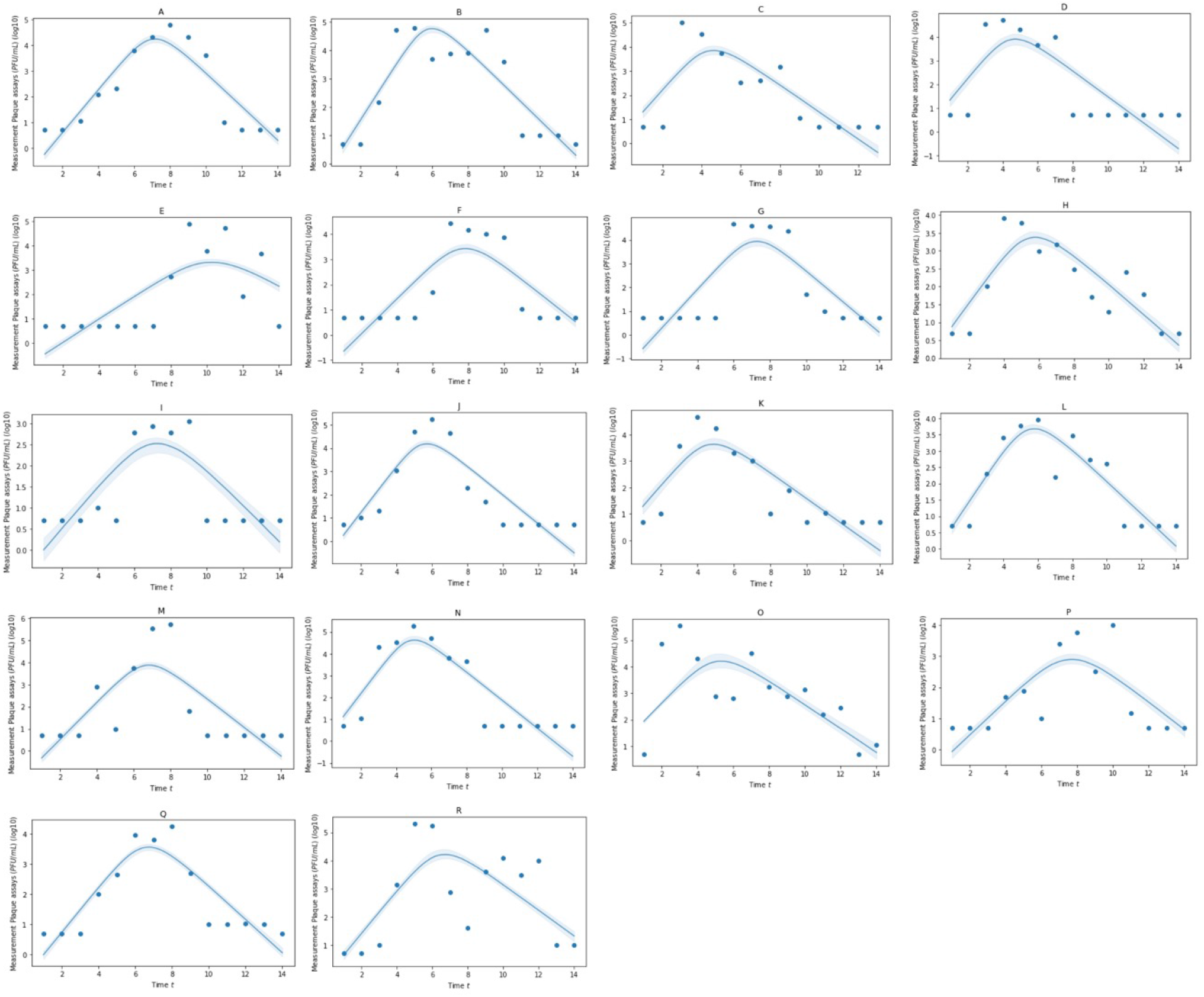
(Scenario 2)Posterior predictions of infectious virus (PFU/mL) from mid-turbinate for 18 participants from the Human Challenge Study Killingley et al. (2022). Model is that derived in equation (2.1) but with values of *τ* greater than 14 days (so there is no immune boost impact from *µ* parameter). Shaded regions represent the 95% credible interval.

**Figure 13:**
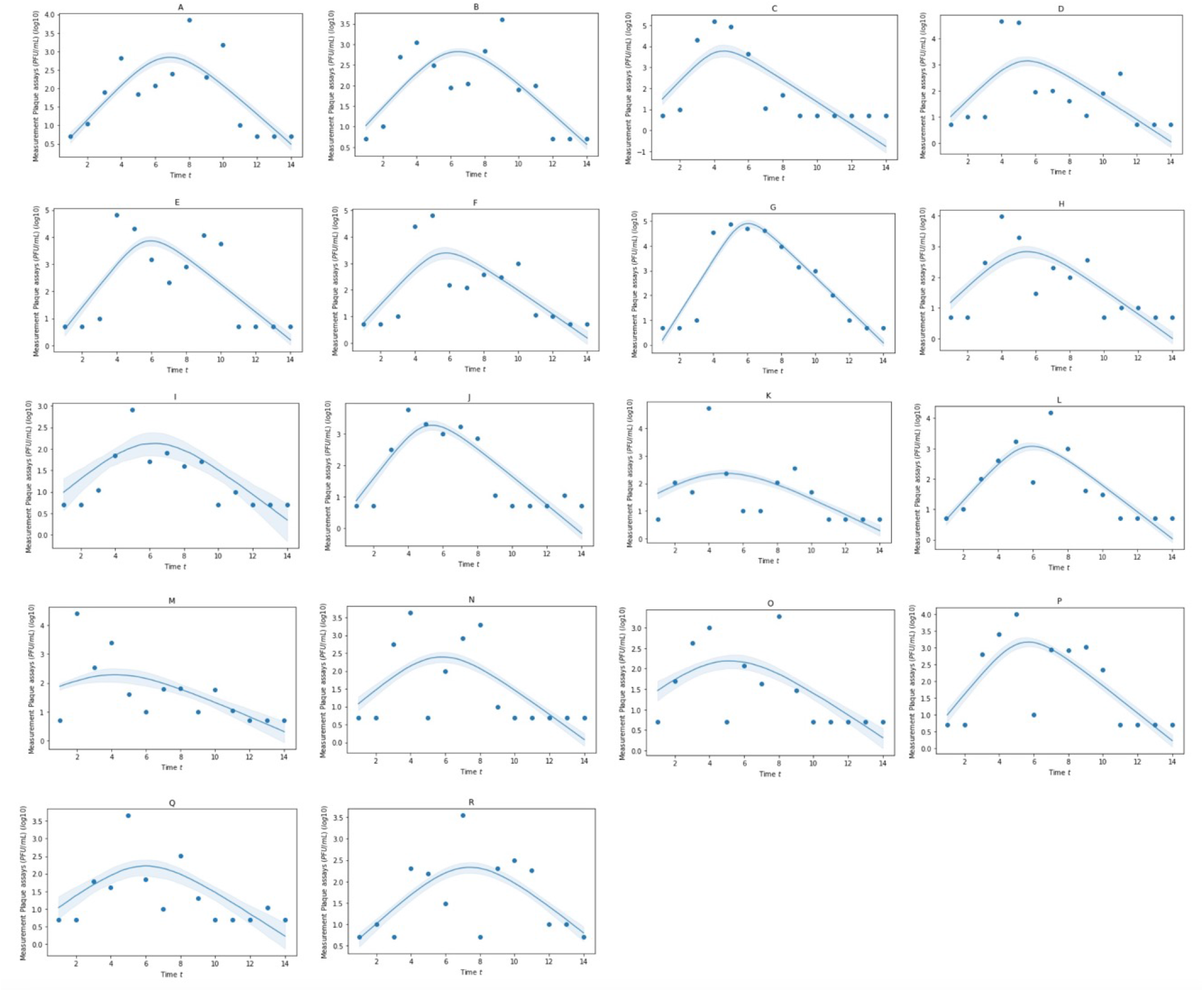
(Scenario 2) Posterior predictions of infectious virus (PFU/mL) from throat for 18 participants from the Human Challenge Study Killingley et al. (2022). Model is that derived in equation (2.1) but with values of *τ* greater than 14 days (so there is no immune boost impact from *µ* parameter). Shaded regions represent the 95% credible interval.

**Figure 14:**
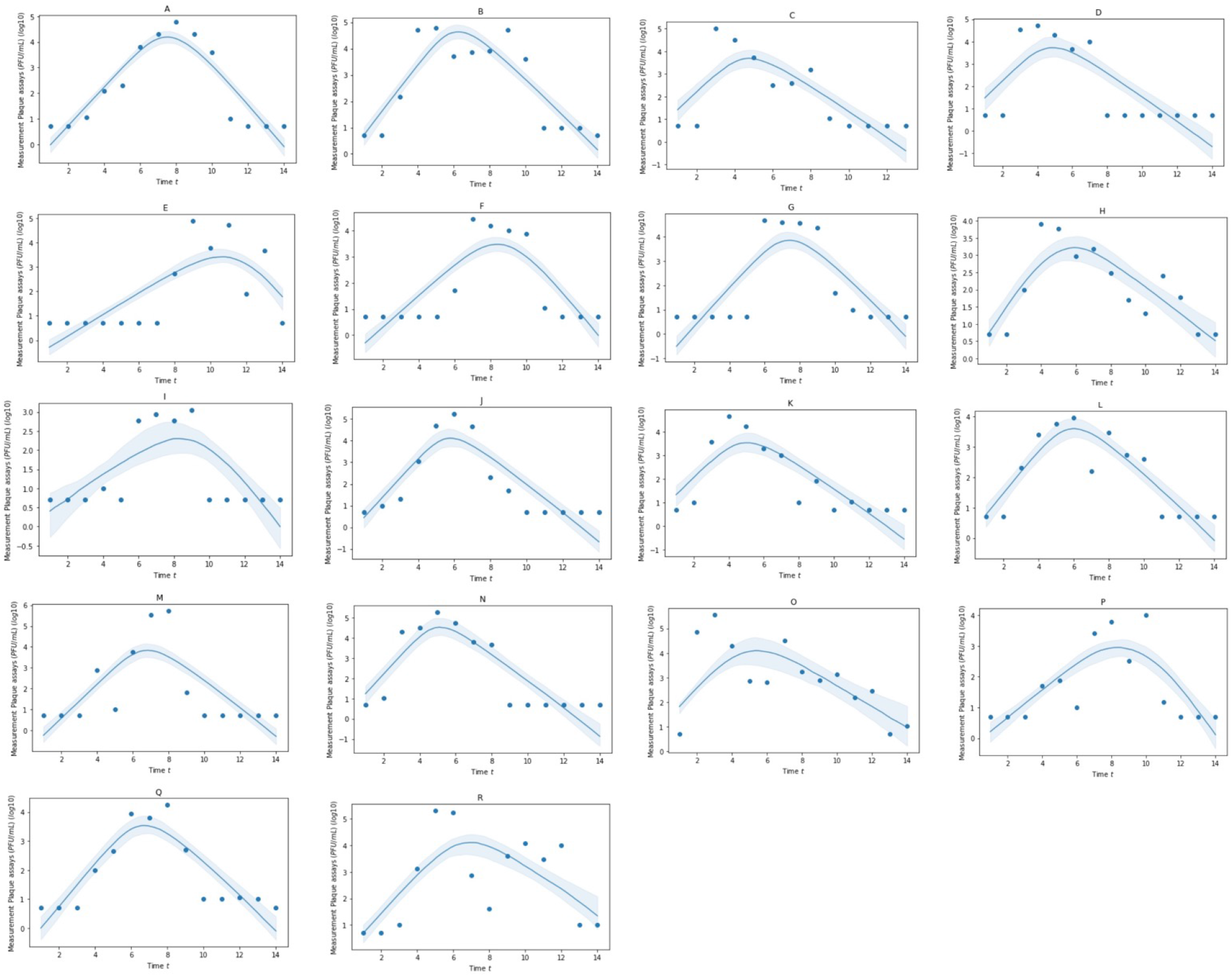
(Scenario 3) Posterior predictions of infectious virus (PFU/mL) from mid-turbinate for 18 participants from the Human Challenge Study Killingley et al. (2022). Model is that derived in equation (2.1). Shaded regions represent the 95% credible interval.

**Figure 15:**
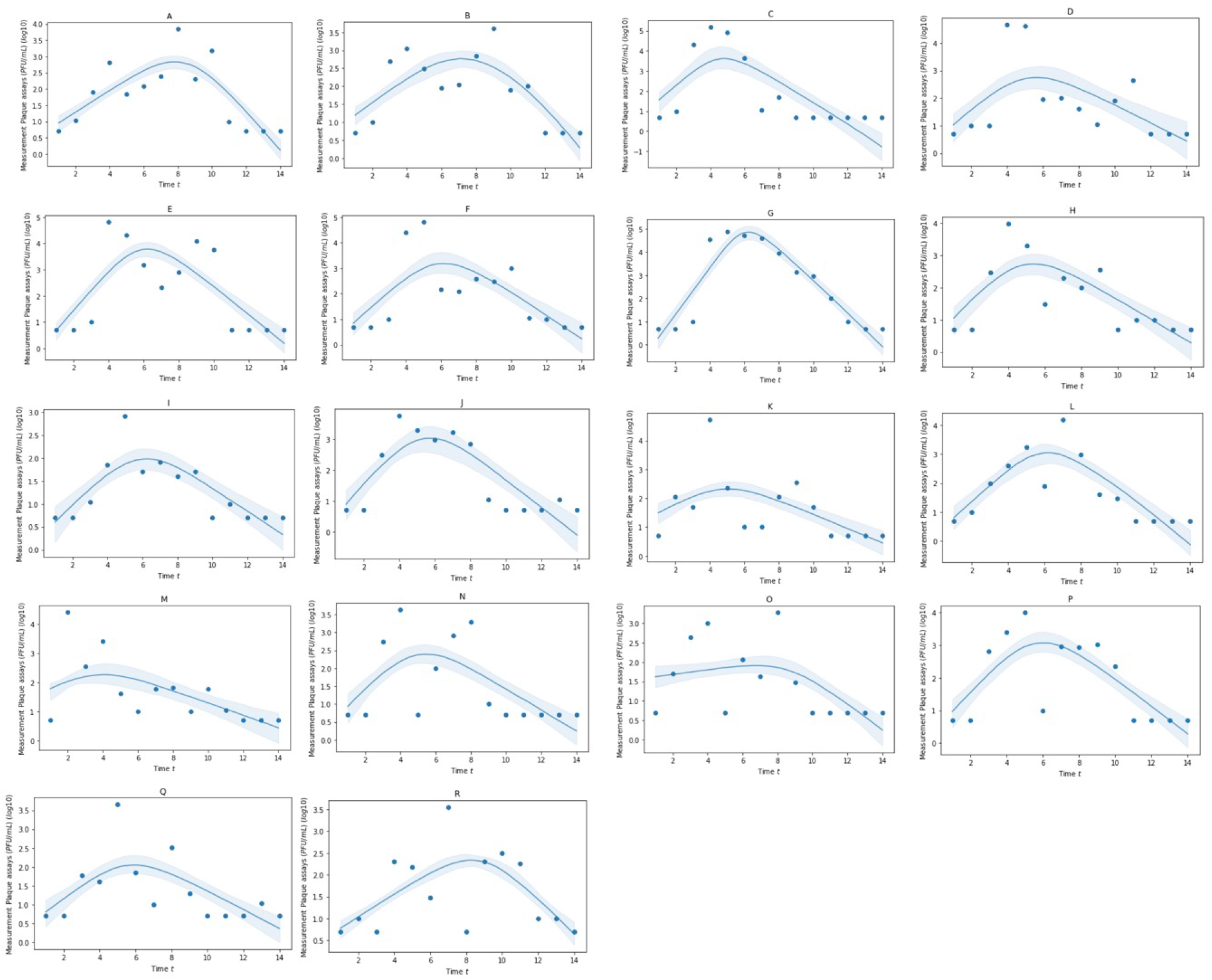
(Scenario 3) Posterior predictions of infectious virus (PFU/mL) from throat for 18 participants from the Human Challenge Study Killingley et al. (2022). Model is that derived in equation (2.1). Shaded regions represent the 95% credible interval.

**Figure 16:**
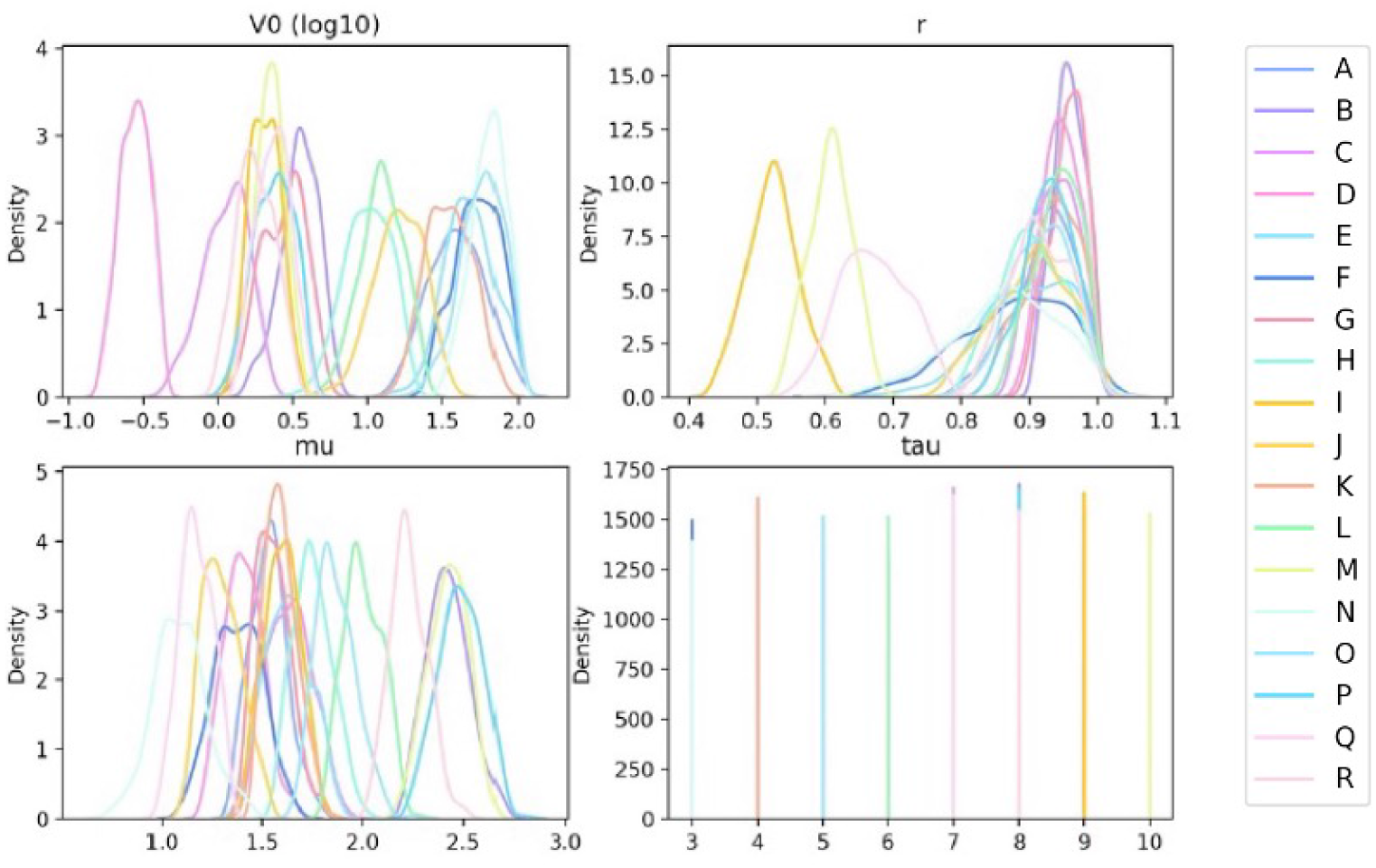
(Scenario 1) Approximate posterior distribution of parameter values from the result of ABC-SMC with model from equation 2.2 using throat data Killingley et al. (2022)

**Figure 17:**
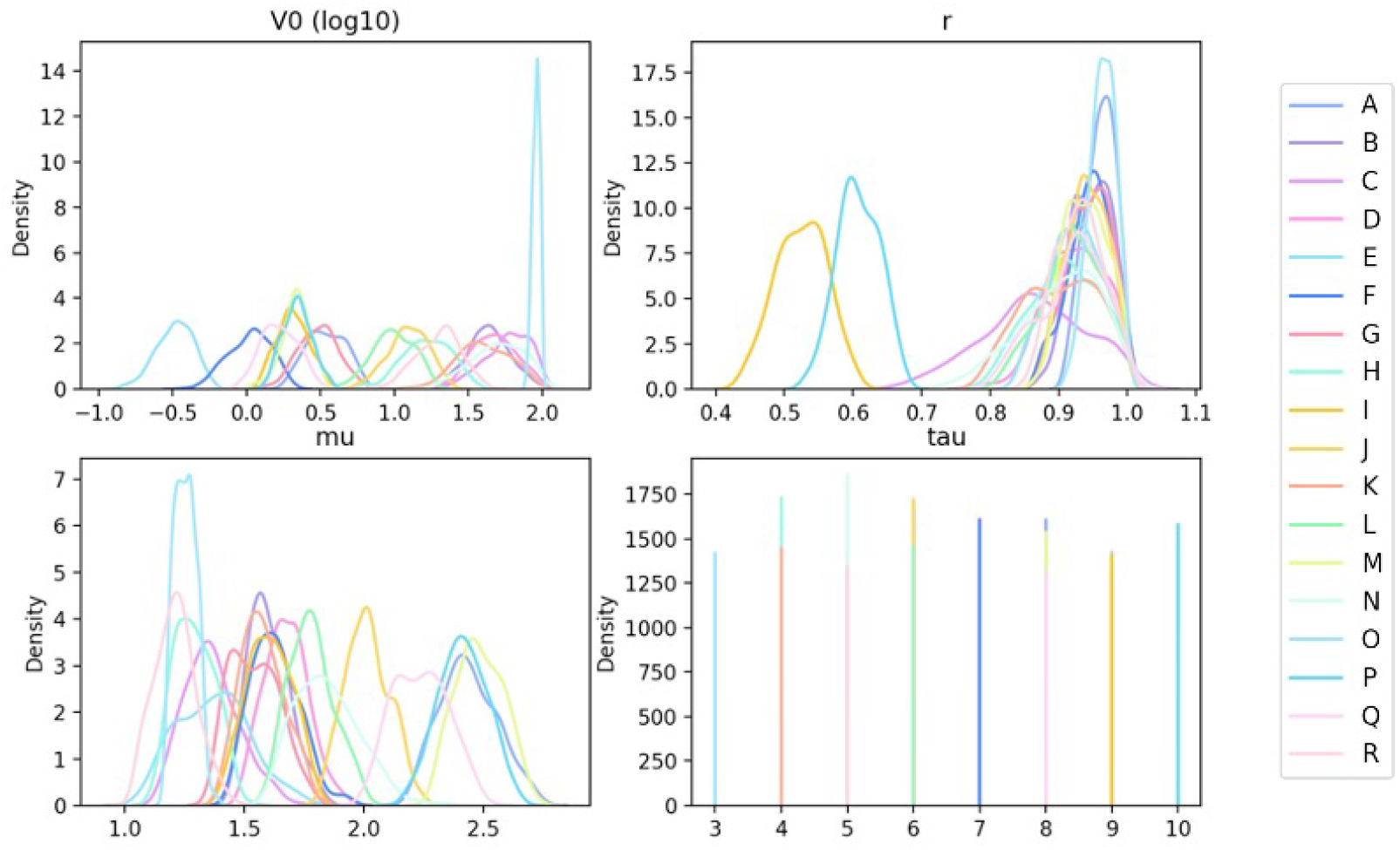
(Scenario 1) Approximate Posterior distribution of parameter values from the result of ABC-SMC with model from equation 2.2 using mid-turbinate data Killingley et al. (2022)

**Figure 18:**
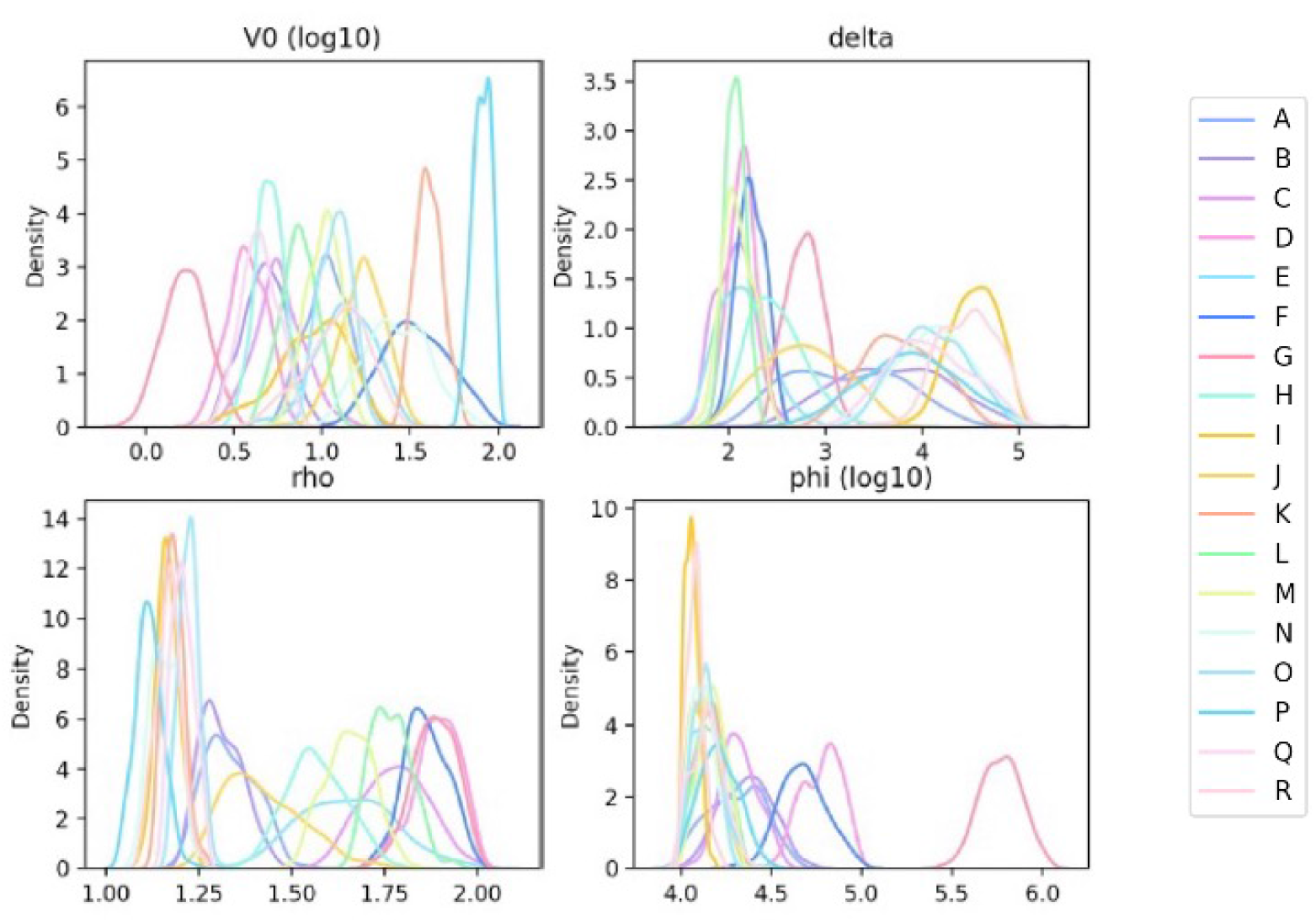
(Scenario 2) Approximate Posterior distribution of parameter values from the result of ABC-SMC with model from equation 2.1 using throat data Killingley et al. (2022)

**Figure 19:**
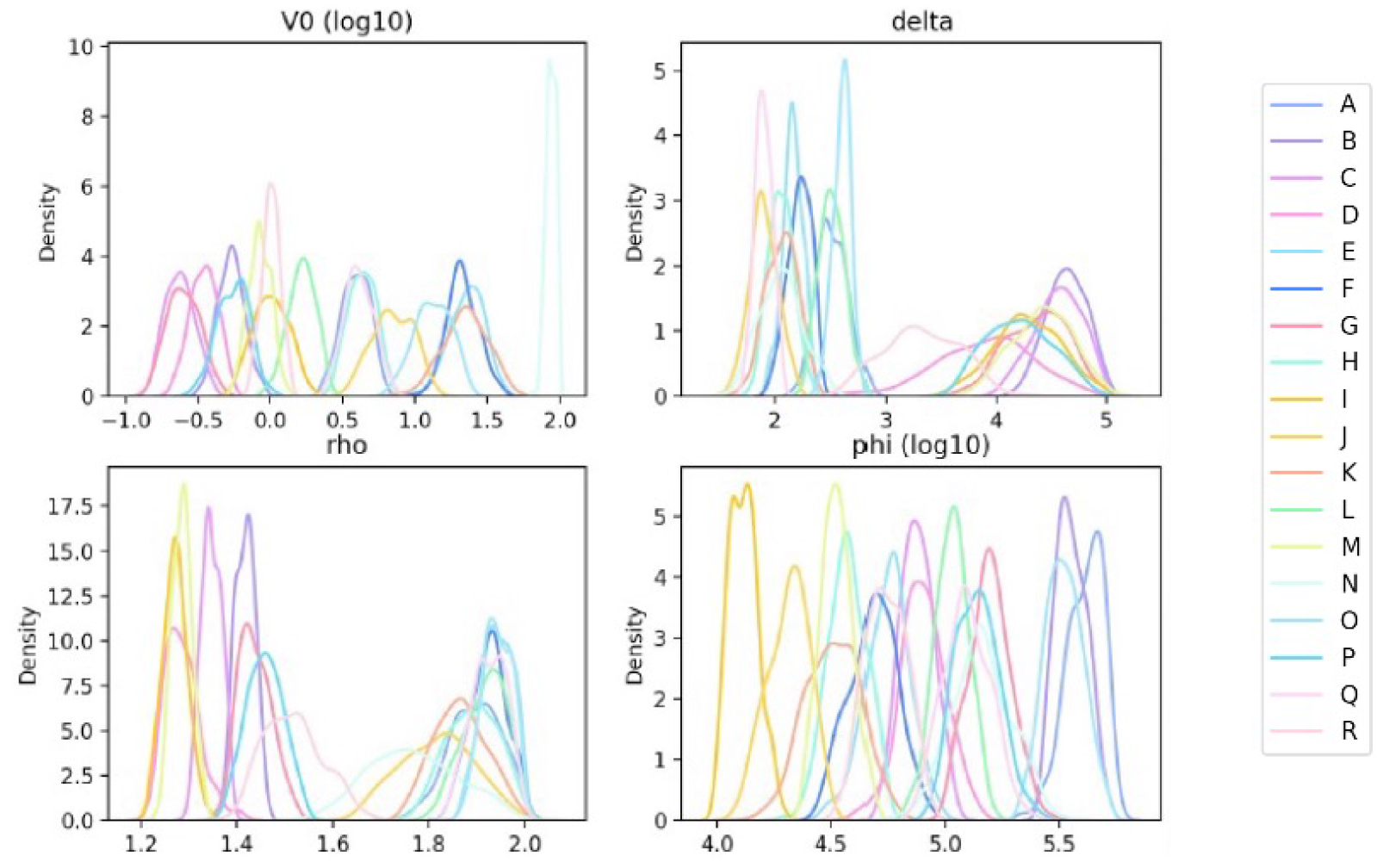
(Scenario 2) Approximate Posterior distribution of parameter values from the result of ABC-SMC with model from equation 2.1 using mid-turbinate data Killingley et al. (2022)

## 5. Discussion

With-in host models for viral loads are helpful for interpolating and extrapolating from experimental data and making predictions for future impacts (such as probability of transmission or effectiveness of testing). A model is essentially a framework for objectively illustrating the consequences of (a chain of) assumptions. These assumptions may be flawed but by having a model one can test hypotheses and potentially show weak assumptions.

The complexity of the immune system and viral dynamics is such that validation of a model is hard in this setting because of individual level variation (so tradition methods of splitting to a training and testing set are not appropriate). Experimental data is expensive to collect and so not always available at scale to calibrate models.

Based upon a range of within-host models used in previous studies, we identify two minimally-complex models, equation 2.1 and equation 2.2, which provides suitable within-host dynamics while keeping a reduced number of parameters to allow inference and limit unidentifiability issues. These two non-dimensional simplified models offer two mechanisms to explain that the decay of viral load within host is caused by the depletion of susceptible cells or the boost of late immunity response respectively. Viral loads measurements from the human challenge study Killingley et al. (2022) have been used in parameter inference and both models could explain the data.

The dose-response models in this paper endow us with more insights into viral load infectiousness. To the best of our knowledge, the extended competing risk framework is first considered in this paper as an functional transform of the viral load to infectiousness. From figure 7-9, we see that the viral load scaling factor plays a critical role in evaluating the probability of infection and smaller values of the scaling factor lead to a narrower infectious period, which indicates that reducing density of viral particles of inhaled air will reduce the chance of infection. Furthermore, compared with the observation of narrow duration, 0.5 to 1 day, of high infectivity in Goyal et al. (2021), the dose-response models in this paper generally exhibit a broader infectious window. For instance, in 7, we see that the exponential function shows a infection window of 6 days. It should be pointed out that the dose-response models used in this paper strictly rely on ID50. Compared with the ID50 from SARS-1, SARS-CoV-2 has much smaller value of ID50, which leads to the significant increase of probability of infection for the same value of viral load and this may explain the superspread of SARS-CoV-2.

This work shows that relatively parsimonious models can be constructed that explain data available. However, two equally valid parameter sets exist that adequately explain data thus from the experimental data alone we cannot infer the key mechanism (depletion of susceptible cells or rapid onset of a boosted immune response). As such mathematical analysis of complex models should be careful of interpretation in case assumptions are flawed. Indeed the argument of model parsimony may not be only decision in model assessment if there is strong virological rationale for mechanisms to be modeled.

## Data Availability

All data produced are available online at https://www.nature.com/articles/s41591-022-01780-9

## Supplementary Material

### S. Model calibration to Human Challenge data

#### S.1. Fast adaptive immune response, large susceptible pool of cells

In this scenario we fit equation (2.2) to the human challenge data Killingley et al. (2022) for mid-turbinate and throat independently. To simplify the model we define *τ* to be the time of the peak viral load and so uncertainty in other parameters may not be fully realised. Figures 10 and 11 show the model fits in both locations for the 18 volunteers.

#### S.2. Slow adaptive immune response, relatively small susceptible pool of cells

In this scenario we fit equation (2.1) to the human challange data but force *τ >* 14 so that neither *τ* or *µ* are estimated. Figures 12 and 13 show the model fits in both locations for the 18 volunteers.

#### S.3. Intermediate adaptive immune response, relatively small susceptible pool of cells

In this scenario we fit equation (2.1) to the human challange data and allow *τ* to be greater than the peak time and up to 14 days. Figures 14 and 15 show the model fits in both locations for the 18 volunteers.

#### S.4. Small susceptible cell population and late immune response boosting

For time *t < τ* then from (2.1) we find

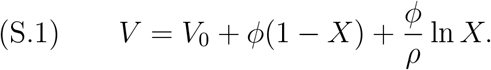

Assuming we have passed the peak viral load (denoted *V*_max_) which occurs when *X* = 1*/ρ* then *ϕ* = (*V*_max_ − *V*_0_)(*ρ*)*/*(*ρ* − 1 − ln (*ρ*)).

We note the equation (2.1) predicts that as *t* → ∞ *V* → 0 and so (S.1) becomes 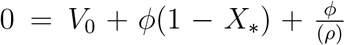 ln *X*_*_ for some *X*_*_ ∈ [0, 1] we can thus define *decay* rates *r*_2_ = *δ*(1 − *ρX*_*_) and *r*_1_ = *r*_2_ + *µ*,

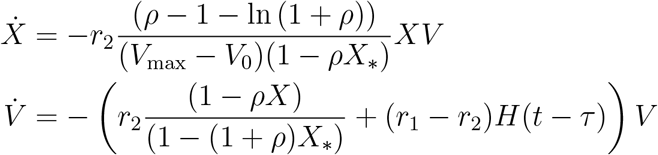

Which though more complex visually may be easier to fit to the data (or at least define meaningful prior estimates for as *V*_max_ is observable statistic, and the decay rates *r*_2_, *r*_1_ and time *τ* can be derived by fitting *V* ∝ exp(−*r*_2_*t* − (*r*_1_ − *r*_2_)*H*(*t* − *τ*)*t*)) to times *t > t*_max_. The parameters *r*_1_ ≥ 0 and *r*_2_ *>* 0 mean that the system grows initially provided *ρ >* 0 (remembering *X*(0) = 1). Note though that this construction of the system requires the solution of the transcendental equation

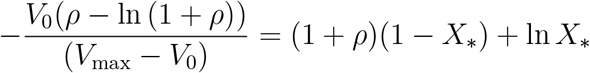

for *X*_*_. Given we expect *V*_max_ ≫ *V*_0_ then *X*_*_ is asymptotically the solution of *ρ* = − ln *X*_*_*/*(1 − *X*_*_) − 1 for a given *ρ* (akin to the usual final size formula from SIR model).

### S. Posterior distribution for individuals

**Figure 20:**
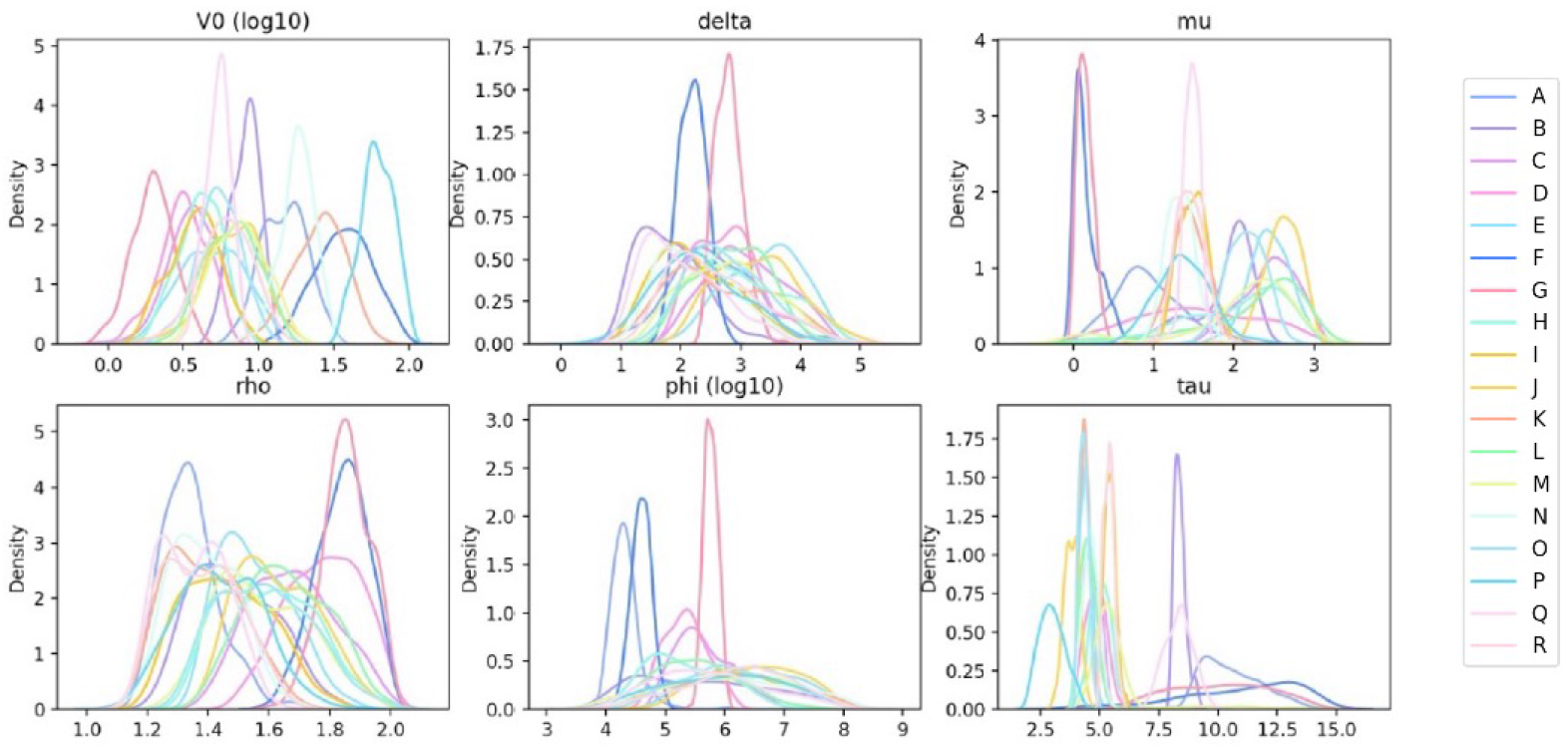
(Scenario 3) Approximate Posterior distribution of parameter values from the result of ABC-SMC with model from equation 2.1 using throat data Killingley et al. (2022)

**Figure 21:**
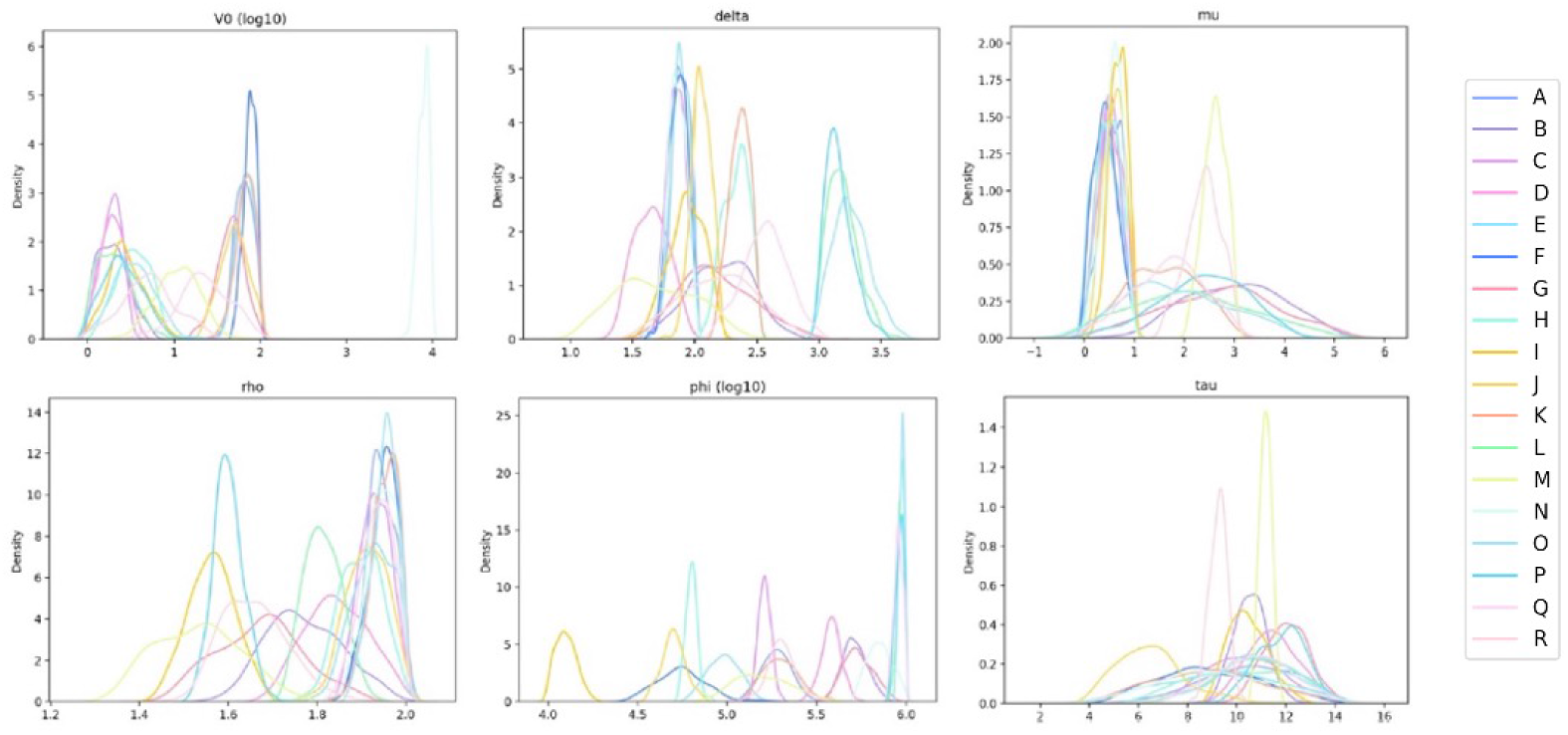
(Scenario 3) Approximate Posterior distribution of parameter values from the result of ABC-SMC with model from equation 2.1 using mid-turbinate data Killingley et al. (2022)

## Notes

### Competing Interest Statement

The authors have declared no competing interest.

### Funding Statement

This report and the research it describes were funded by the PROTECT COVID-19 National Core Study on transmission and environment, which is managed by the Health and Safety Executive (HSE) on behalf of HM Government. Its contents, including any opinions and/or conclusions expressed, are those of the authors alone and do not necessarily reflect UK Government or HSE policy.

